# Gut microbiota impacts bone via *B.vulgatus*-valeric acid-related pathways

**DOI:** 10.1101/2020.03.16.20037077

**Authors:** Xu Lin, Hong-Mei Xiao, Hui-Min Liu, Wan-Qiang Lv, Jonathan Greenbaum, Si-Jie Yuan, Rui Gong, Qiang Zhang, Yuan-Cheng Chen, Cheng Peng, Xue-Juan Xu, Dao-Yan Pan, Zhi Chen, Zhang-Fang Li, Rou Zhou, Xia-Fang Wang, Jun-Min Lu, Zeng-Xin Ao, Yu-Qian Song, Yin-Hua Zhang, Kuan-Jui Su, Xiang-He Meng, Chang-Li Ge, Feng-Ye Lv, Xing-Ming Shi, Qi Zhao, Bo-Yi Guo, Neng-Jun Yi, Hui Shen, Christopher J. Papasian, Jie Shen, Hong-Wen Deng

**Affiliations:** Tulane Center for Bioinformatics and Genomics, School of Public Health and Tropical Medicine, Tulane University, New Orleans, LA 70112, USA; Department of Endocrinology and Metabolism, The Third Affiliated Hospital of Southern Medical University, Guangzhou 510630, China; Center of System Biology, Data Information and Reproductive Health, School of Basic Medical Science, Central South University, Changsha 410008, China; LC-Bio Technologies (Hangzhou) CO., LTD., Hangzhou 310018, China; Departments of Neuroscience & Regenerative Medicine and Orthopaedic Surgery, Medical College of Georgia, Augusta University, Augusta, GA, USA; Department of Preventive Medicine, College of Medicine, University of Tennessee Health Science Center, Memphis, TN 38163, USA; Department of Biostatistics, University of Alabama at Birmingham, Alabama 35294, USA; Department of Basic Medical Science, School of Medicine, University of Missouri-Kansas City, 2411 Holmes Street, Kansas City, MO 64108; Shunde Hospital of Southern Medical University (The First People’s Hospital of Shunde), No.1 of Jiazi Road, Lunjiao, Shunde District, Foshan City, Guangdong Province, China

## Abstract

Although gut microbiota influences osteoporosis risk, the individual species involved, and underlying mechanisms, are unknown. We performed integrative analyses in a Chinese cohort with metagenomics/targeted metabolomics/whole-genome sequencing. *Bacteroides vulgatus* was found negatively associated with bone mineral density (BMD), this association was validated in US Caucasians. Serum valeric acid was positively associated with BMD, and *B.vulgatus* causally downregulated it. Ovariectomized mice fed *B.vulgatus* had decreased bone formation and increased bone resorption, lower BMD and poorer bone micro-structure. Valeric acid suppressed NF-κB p65 protein production (pro-inflammatory), and enhanced IL-10 mRNA expression (anti-inflammatory), leading to suppressed maturation of osteoclast-like cells, and enhanced maturation of osteoblasts *in vitro. B.vulgatus* and valeric acid represent promising targets for osteoporosis prevention/treatment.

---

Osteoporosis (OP), mainly characterized by low bone mineral density (BMD), predisposes to osteoporotic fracture, and is most prevalent in postmenopausal women (termed postmenopausal osteoporosis (PMOP)). 4.9 million post-menopausal women in US had osteoporotic fractures between 2000 and 2011, with greater hospital costs than myocardial infarction, stroke, and breast cancer ^1,2^. The pathogenic mechanisms for PMOP and OP remain unclear, and current intervention and treatment options are not satisfactory ^1,2^.

Human studies have focused on the potential impact of inherent (e.g., (epi-)genome, transcriptome, proteome, metabolome) and external (e.g., environmental, medical/medication/nutrition, lifestyle) risk factors for OP ^3^. Recently, however, experimental animal models have identified strong associations between gut microbiota (GM) and bone ^4^. For example, Sjogren *et al*. found that absence of GM in mice leads to increased bone mass that normalizes following GM colonization ^5^. In humans, four studies suggested the potential importance of GM for bone health ^6-9^, however, these studies had small sample sizes and did not identify specific bacterial species and associated functional mechanisms.

To identify specific significant gut bacterial species and their functional mechanisms on bone health, we performed a novel systematic integrative multi-omics analysis of the human genome, GM and targeted metabolome [serum short chain fatty acids (SCFAs)] in 517 peri- and post-menopausal Chinese women. We identified several bacterial species SCFAs and functional pathways that significantly impacted BMD. We validated the major results in US Caucasians and investigated causal roles of individual bacterial species and SCFAs in regulating bone metabolism *in vitro* in cells and *in vivo* in mice. Our work is the first to identify individual BMD-associated GM species and particularly GM-derived functional metabolites in humans, and establish specific pathogenic mechanisms by which GM influences OP risk. Our results lay a foundation for potential prevention/intervention/treatment of OP through GM and metabolome alteration.

## Chinese study cohort characteristics

517 peri-/post-menopausal Chinese women were randomly recruited from Guangzhou City, China; 84% were post-menopausal and 16% peri-menopausal, based on years since menopause (YSM) ^10^. Mean YSM and follicle stimulating hormone (FSH) levels were 1.96 years and 76.24 mIU/mL, respectively (Table 1). Age (mean ± standard deviation [SD], 52.85 ± 2.92 years) and body mass index (BMI) (22.97 ± 2.87 kg/m^2^) were relatively homogeneous. Mean BMD of the lumbar spine (L1-L4), left total hip (HTOT, including femoral neck, trochanter, and inter-trochanter), femoral neck (FN), and ultra-distal radius and ulna (UD-RU) were 1.05 g/cm^2^, 0.93 g/cm^2^, 0.86 g/cm^2^, and 0.40 g/cm^2^, respectively. Based on WHO criteria, 54.5% of the subjects had normal BMD (T-score ≥ -1), 38.5% osteopenia (-2.5 < T-score < -1), and 7% OP (T-score ≤ -2.5). Lifestyle factors (e.g., alcohol consumption, smoking, calcium supplementation, exercise) and socioeconomic status (e.g., education and family annual income) are shown in Extended Data (ED) Table 1. Stool DNA and blood samples were collected for metagenomic, genomic, and metabolomic analyses.

**Table 1.**
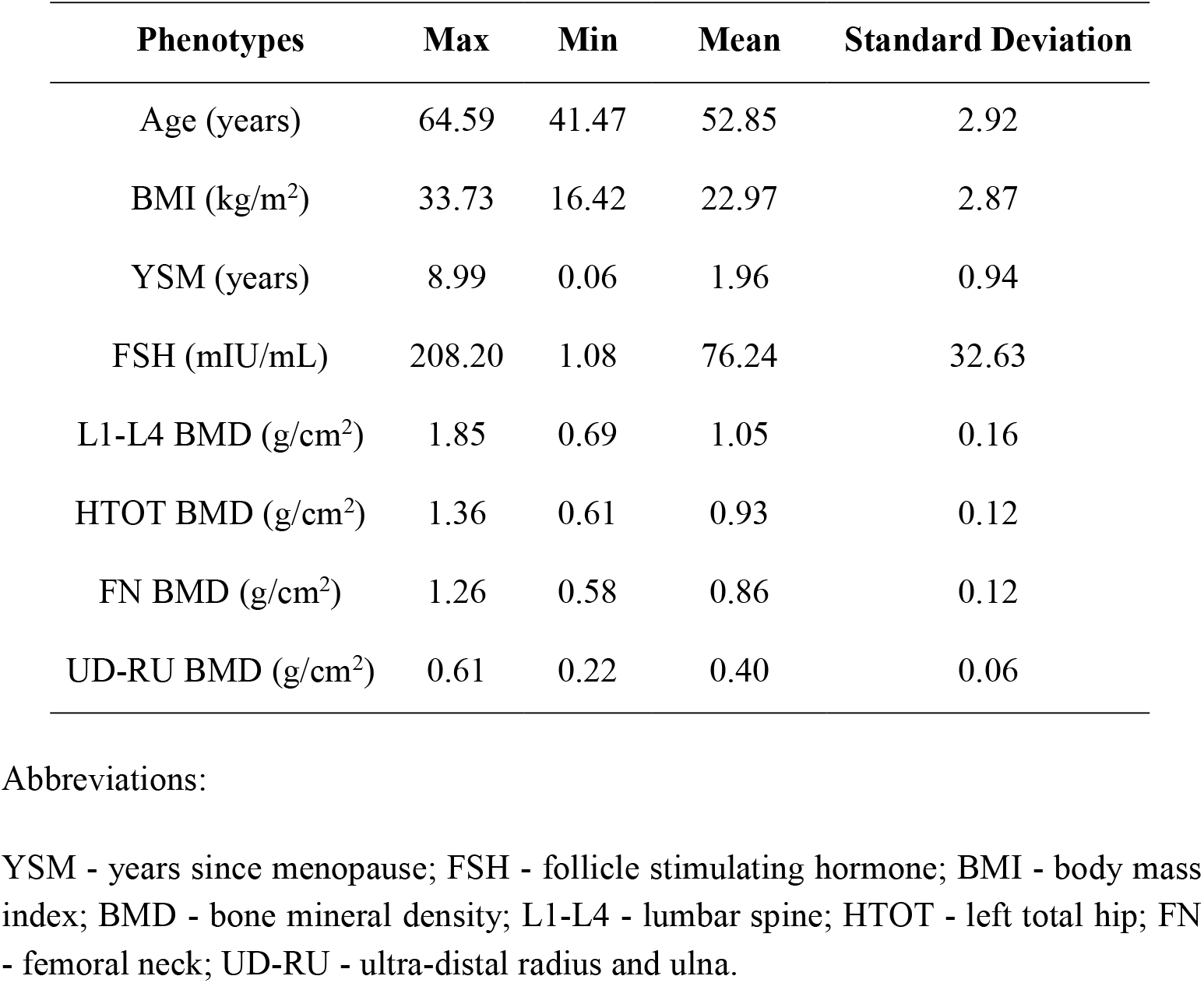
Characteristics of the Chinese study cohort

| Phenotypes | Max | Min | Mean | Standard Deviation |
| --- | --- | --- | --- | --- |
| Age (years) | 64.59 | 41.47 | 52.85 | 2.92 |
| BMI (kg/m<sup>2</sup>) | 33.73 | 16.42 | 22.97 | 2.87 |
| YSM (years) | 8.99 | 0.06 | 1.96 | 0.94 |
| FSH (mIU/mL) | 208.20 | 1.08 | 76.24 | 32.63 |
| L1-L4 BMD (g/cm<sup>2</sup>) | 1.85 | 0.69 | 1.05 | 0.16 |
| HTOT BMD (g/cm<sup>2</sup>) | 1.36 | 0.61 | 0.93 | 0.12 |
| FN BMD (g/cm<sup>2</sup>) | 1.26 | 0.58 | 0.86 | 0.12 |
| UD-RU BMD (g/cm<sup>2</sup>) | 0.61 | 0.22 | 0.40 | 0.06 |

3 YSM - years since menopause; FSH - follicle stimulating hormone; BMI - body mass 4 index; BMD - bone mineral density; L1-L4 - lumbar spine; HTOT - left total hip; FN 5 - femoral neck; UD-RU - ultra-distal radius and ulna.

## Correlation between GM and BMD

We performed shotgun metagenomics sequencing on stool DNA samples and obtained ∼7.35 Gbp/sample sequencing data on average. We calculated unigene (non-redundant genes) counts at different sample sizes. The number of unigenes increased with sample size, approaching a plateau (exceeded 7 × 10^6^) with our study sample (Figure 1a), indicating that our sample size was appropriate, large and reasonably powerful for this investigation.

**Figure 1.**
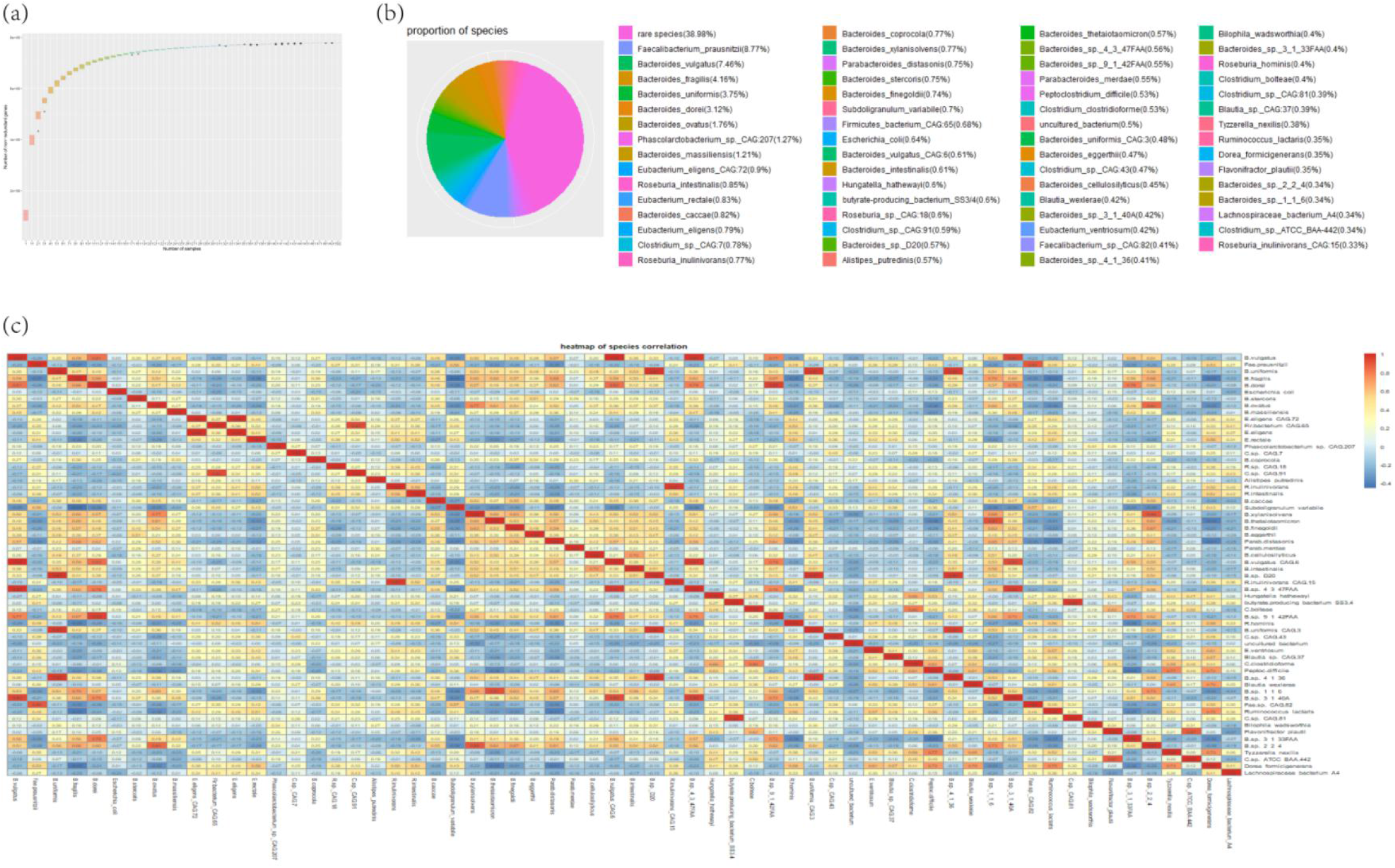
Unigene accumulation curve, bacterial species composition and correlation. (a) Unigene accumulation curve: the number of unigenes (y-axis) was plotted against the number of samples (x-axis). (b) Bacterial species composition in the study cohort composed of rare and non-rare bacterial species. Different colours represent different bacterial species. Numbers in parentheses indicate the proportion of each bacterial species. “Rare species” include all those bacterial species with relative abundances less than 0.10%. (c) Heatmap of co-occurrence between the relative abundances of non-rare species. Numbers in the plot represent Spearman correlation coefficients of pairwise two bacterial species.

We identified 10,303 species by taxonomy annotation (ftp://ftp.ncbi.nlm.nih.gov/blast/db/FASTA/nr.gz). 62 species were non-rare (relative abundance > 0.10%) ^11^ accounting for 61.02% of the whole GM. The rest were rare species (Figure 1b). The 3 most common species were *Faecalibacterium prausnitzii* (8.77%), *Bacteroides vulgatus* (7.46%), and *Bacteroides fragilis* (4.16%). 26 and 27 of the non-rare species had high positive (Spearman correlation coefficients [γ’s] > 0.80, *p*-values < 0.001) and negative (γ’s < -0.30, *p*-values < 0.001) correlations, respectively, with one another (Figure 1c). This suggests that interactions between GM species promote co-existence (when γ is > 0.80) or suppression (when γ is < -0.30) of each other.

We used “vegan” package and Microbiome Regression-based Kernel Association Test (MiRKAT) ^12^ to calculate α-diversity (Shannon index) and β-diversity (optimal kernel, based on weighted and unweighted UniFrac distance matrices and Bray-Curtis distance), and evaluated their associations with BMD. GM biodiversity was negatively associated with forearm BMD (ED Table 2), including ultra-distal radius BMD (UD-R BMD, partial γ = -0.081, *p*-value = 0.072, *p*-value of MiRKAT = 0.005), ultra-distal ulna BMD (UD-U BMD, partial γ = -0.098, *p*-value = 0.029, *p*-value of MiRKAT = 0.014), and UD-RU BMD (partial γ = -0.097, *p*-value = 0.031, *p*-value of MiRKAT = 0.010). The *p*-value significance remained when adjusted with false discovery rate for multiple testing (*q*-values ∼0.10 or less). Although no significant association was found between GM biodiversity and BMD at L1-L4 or HTOT, we hypothesized that individual bacterial species contributed to these phenotypes.

**Table 2.** Correlation between GM functional capacity and BMD variation

| Module ID | $\gamma$ | *p*-value | *Q*-value | Module names | Phenotypes |
| --- | --- | --- | --- | --- | --- |
| M00002 | -0.122 | 0.007 | 0.234 | Glycolysis | L1-L4 BMD |
| M00048 | -0.119 | 0.008 | 0.234 | IMP biosynthesis | L1-L4 BMD |
| M00855 | -0.119 | 0.008 | 0.234 | Glycogen degradation | L1-L4 BMD |
| M00002 | -0.118 | 0.009 | 0.153 | Glycolysis | UD-RU BMD |
| M00003 | -0.119 | 0.008 | 0.153 | Gluconeogenesis | UD-RU BMD |
| M00050 | -0.143 | 0.002 | 0.079 | GTP biosynthesis | UD-RU BMD |
| M00727 | -0.135 | 0.003 | 0.079 | CAMP resistance | UD-RU BMD |

Note:
$\gamma$ - coefficient of the partial Spearman correlation between GM functional module and phenotype; *p*-value - *p*-value of the coefficient in partial Spearman correlation analysis; *Q*-value - *p*-value adjusted by false discovery rate; Glycolysis - core module involving three-carbon compounds; IMP biosynthesis - PRPP + glutamine → IMP; Glycogen degradation - glycogen → glucose-6P; Gluconeogenesis - oxaloacetate → fructose-6P; GTP biosynthesis - IMP → GDP, GTP; CAMP resistance - N-acetylmuramoyl-L-alanine amidase AmiA and AmiC. We only tested the modules whose relative abundance > 0.10%.
GM - gut microbiota; BMD - bone mineral density; L1-L4 - lumbar spine; UD-RU - ultra-distal radius and ulna; PRPP - phosphoribosylpyrophosphate; IMP - inosine monophosphate; GDP - guanosine 5'-diphosphate; GTP - guanosine 5'-triphosphate; CAMP - cationic antimicrobial peptide.

We performed constrained linear regression analysis ^13^, and found several individual species significantly associated with BMD (ED Table 3). Specifically, *Bacteroides vulgatus* (regression coefficient [β] = -0.027, *p*-value = 0.032), *Bacteroides thetaiotaomicron* (β = 0.027, *p*-value = 0.021), *Butyrate-producing bacterium_SS3/4* (β = -0.024, *p*-value = 0.040), *Clostridium bolteae* (β = 0.045, *p*-value = 0.036), and *Bacteroides sp_9_1_42faa* (β = 0.033, *p*-value = 0.031) were associated with L1-L4 BMD. Additionally, *Butyrate-producing bacterium_SS3/4* was associated with HTOT BMD (β = -0.022, *p*-value = 0.009) and FN BMD (β = -0.021, *p*-value = 0.014); *Bacteroides ovatus* was associated with UD-RU BMD (β = 0.011, *p*-value = 0.046); and *Bacteroides intestinalis* was associated with HTOT BMD (β = -0.021, *p*-value = 0.043).

After function annotation and obtaining KEGG modules, we performed partial Spearman correlation analysis to identify relationships between GM functional capacity and BMD variation. Three functional modules were significantly negatively associated with L1-L4 BMD and four modules were negatively associated with UD-RU BMD (γ’s < -0.10, *p*-values < 0.010) (Table 2). For example, for L1-L4 BMD, significantly associated modules included “M00002: glycolysis, core module involving three-carbon compounds”, “M00048: inosine monophosphate biosynthesis, phosphoribosylpyrophosphate + glutamine → inosine monophosphate”, and “M00855: glycogen degradation, glycogen → glucose-6P”. All these modules involve pathways whereby GM produces metabolites from ingested foods, suggesting that these metabolites influence BMD.

## SCFAs significantly associated with BMD

GM may affect host health through SCFAs ^14^. Therefore, we analyzed serum SCFA levels by targeted metabolomics. By multiple linear regression analysis, valeric acid was positively associated with L1-L4 BMD (β = 0.040, *p*-value = 0.029). The results (non-significant) of other kinds of SCFAs (including caproic acid, isovaleric acid, butyric acid, acetic acid, and isobutyric acid) were shown in ED Table 4. Furthermore, five bacterial species were significantly associated with valeric acid (ED Table 5). *Alistipes putredinis, Bacteroides caccae*, and *Bacteroides cellulosilyticus* were positively associated with valeric acid (β’s > 0.05, *p*-values < 0.05, ED Table 5). *B.vulgatus* and *B.intestinalis*, which were negatively associated with L1-L4 and HTOT BMDs (ED Table 3), were also negatively associated with valeric acid (β’s < -0.10, *p*-values < 0.01, ED Table 5). Since only *B.vulgatus* was significantly associated with both L1-L4 BMD and valeric acid, we subsequently focused on this species.

Since valeric acid is produced by probiotic bacteria ^15^, we calculated the Spearman correlation between *B.vulgatus* and common probiotic bacteria ^16, 17^. The relative abundance of *B.vulgatus* was negatively associated (γ = -0.24, *p*-value = 8.3 × 10^−8^) with the sum of relative abundance of common probiotic bacteria (each still rare (< 0.10%) so pooled for analyses). We used Mendelian randomization (MR) approach ^18^ to assess the potential causality of *B.vulgatus* (as exposure) on valeric acid (as outcome), using their associated SNPs derived from whole genome sequencing for human subjects as instrumental variables. Most MR methods, including the weighted median method, maximum likelihood estimation, and inverse-variance weighted indicated that *B.vulgatus* may causally down-regulate valeric acid (β’s < -0.07, *p*-values < 0.05) (ED Table 6). MR-Egger (intercept) result (*p*-value = 0.517) showed no horizontal pleiotropy for *B.vulgatus* to influence valeric acid levels.

## *B.vulgatus*/BMD associations in US Caucasians

To assess the robustness of our major findings, we tested associations between *B.vulgatus* and human BMD in an independent cohort of 59 post-menopausal Caucasian females, aged ≥ 60 years, in New Orleans, Louisiana. Mean (SD) age and BMI were 66.98 (5.65) years and 27.84 (8.50) kg/m^2^, respectively. Mean BMD for L1-L4, HTOT, FN, UD-R, and UD-U were 0.92 g/cm^2^, 0.80 g/cm^2^, 0.67 g/cm^2^, 0.37 g/cm^2^, and 0.28 g/cm^2^, respectively. Lifestyle information is shown in ED Table 7. We detected an association between *B.vulgatus* and HTOT BMD (β = -0.018, *p*-value = 0.029), supporting *B.vulgatus*’s association with BMD in ethnically distinct populations.

## *In vivo* and *in vitro* validation

To evaluate potential causal effects of *B.vulgatus* on bone metabolism (focusing on lumbar vertebrae), we ovariaectomized (OVX) 8-week-old female C57BL/6 mice (n = 18) to model post-menopause, and performed oral gavage experiments. After gavaging with *B.vulgatus*/normal saline (NS) for eight weeks after OVX, we found lower BMD (*p*-value < 0.05, Figure 2a), increased disconnection and separation of the trabecular bone network, and reduction of trabecular number (Figure 2d and Figure 2e) in the *B.vulgatus* treated group. The *B.vulgatus* treated group also showed lower (two-sample t-test, *p*-value < 0.05) bone formation levels (measured by serum osteocalcin, Figure 2b) and higher (two-sample t-test, *p*-value < 0.05) bone resorption levels (measured by type I collagen serum C-telopeptide, Figure 2c). This demonstrates that *B.vulgatus* suppressed bone formation and promoted bone resorption leading to reduced lumbar vertebral BMD and deteriorated bone architecture.

**Figure 2.**
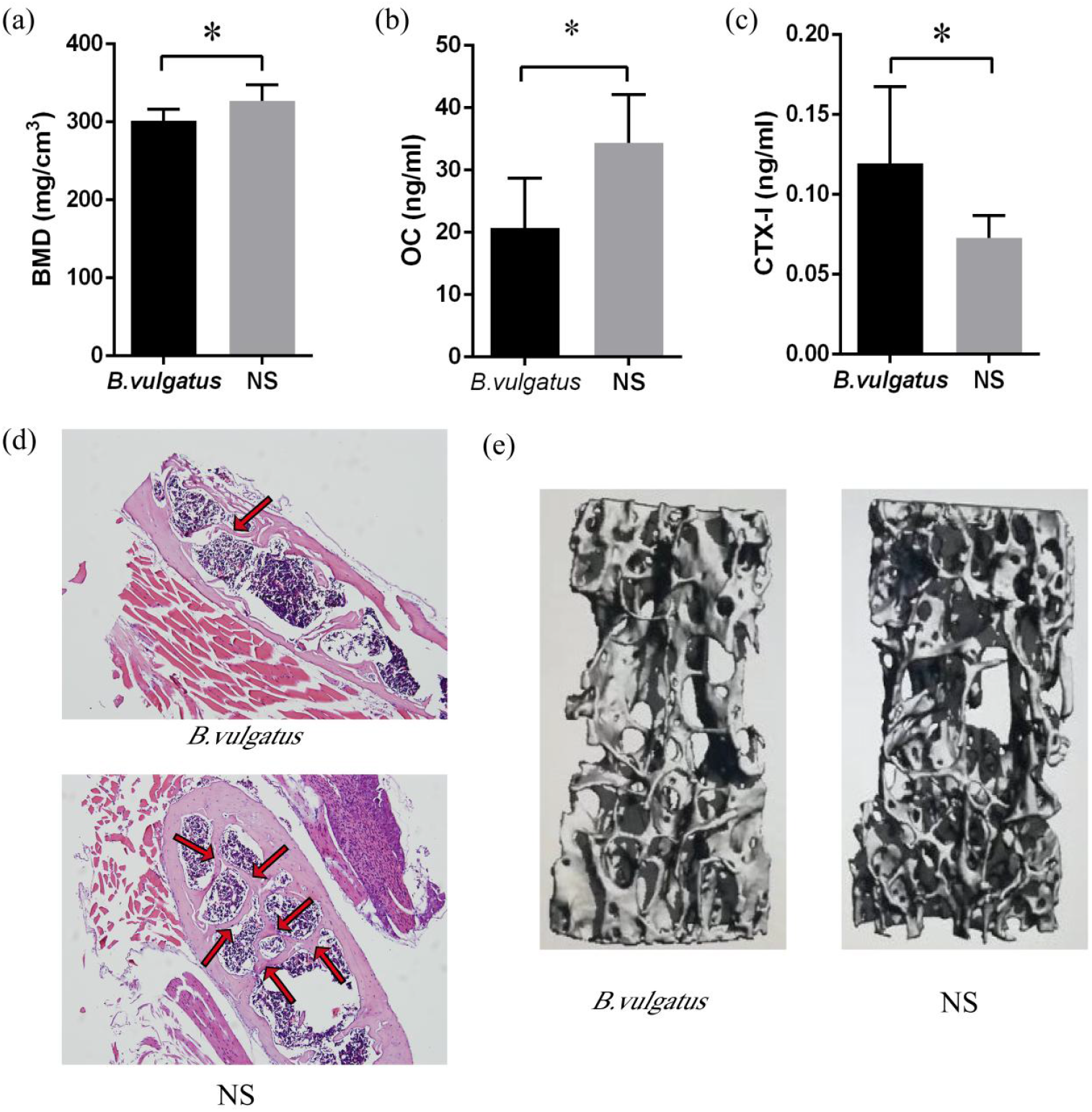
*Bacteroides vulgatus* regulates bone remodeling in mice *in vivo*. Changes in various bone-associated phenotypes in mice gavaged with *B. vulgatus* or normal saline (NS) for 8 weeks (performed at age of 8 weeks after ovariectomy): (a) bone mineral density (BMD) of the 5^th^ lumbar vertebral body; (b) serum osteocalcin (OC) concentrations; (c) serum C-telopeptide of type I collagen (CTX-I) concentrations; (d) representative histomorphology of the 5^th^ lumbar vertebral body using hematoxylin-eosin staining. Red arrows point to trabecular bone; (e) representative micro-computed tomograph images of the 5^th^ lumbar vertebral body by three-dimensional reconstructions. (Note: in A, B, and C, n = 18, and _*_ indicates *p*-value < 0.05.)

Since our results suggested that *B.vulgatus* affects BMD by down-regulating valeric acid production, we investigated the effects of valeric acid on osteoclast and osteoblast differentiation *in vitro*. Murine macrophages (RAW264.7) and pre-osteoblasts (MC3T3-E1) were differentiated with/without valeric acid. After inducing osteoclast differentiation of RAW264.7 cells with RANKL (50 ng/mL), 5 days treatment with valeric acid (1 mM) significantly decreased the proportion of mature osteoclast-like cells (TRAP positive and with ≥ 3 nuclei) vs. controls (Figure 3a), indicating that valeric acid significantly inhibited osteoclast differentiation. Treatment with valeric acid (1 mM) for 14 days also significantly increased differentiation of MC3T3-E1 into osteoblasts, and mineralization of the extracellular matrix (Figure 3b). Therefore, valeric acid inhibited differentiation of osteoclast-like cells, and promoted differentiation of, and mineralization by, osteoblasts.

**Figure 3.**
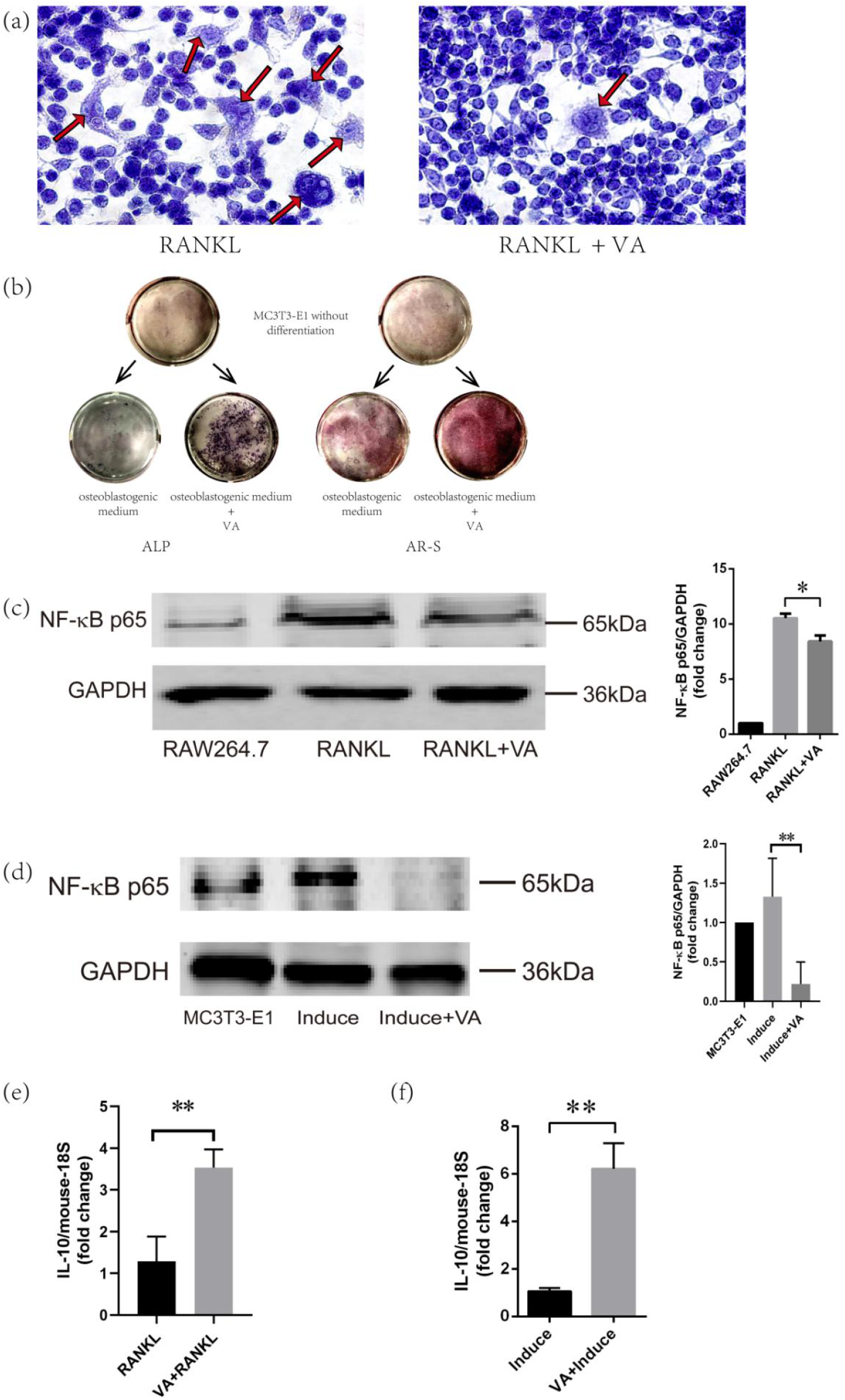
Valeric acid influences osteoclast and osteoblast differentiation *in vitro*. Effects of valeric acid (VA) on osteoclast differentiation of RAW264.7 cells (induced by receptor activator of nuclear factor-κB ligand [RANKL]) and osteoblast differentiation of MC3T3-E1 cells: (a) Microscopic images of tartrate-resistant acid phosphatase staining of osteoclast-like cells induced from RAW264.7 cells after 5 days of osteoclastogenesis with/without VA treatment. Red arrows indicate osteoclast-like cells. (b) Alkaline phosphatase (ALP) staining for osteoblast differentiation and alizarin red S (AR-S) staining for extracellular matrix mineralization by MC3T3-E1 cells after 14 days of osteoblastogenesis with/without VA treatment; (c) Western blot of NF-κB p65 protein in osteoclast-like cells with/without VA treatment; (d) Western blot of NF-κB p65 protein in osteoblasts with/without VA treatment; the “induce” means the MC3T3-E1 cells were induced into osteoblasts by osteoblastogenic medium without VA; (e) IL-10 mRNA expression in osteoclast-like cells with/without VA; (f) IL-10 mRNA expression in osteoblasts with/without VA. _*_ indicates *p*-value < 0.05, and _**_ means *p*-value < 0.01.

Because inflammatory pathways play important roles in bone metabolism ^19^ and GM may influence bone metabolism through its effects on inflammation ^4^, we measured expression levels of several critical inflammatory genes (e.g., NF-κB p65 and IL-10) in cultured osteoclast-like cells and osteoblasts. After treating osteoclast-like cells and osteoblasts with valeric acid for 5 and 14 days, respectively, IL-10 (anti-inflammatory cytokine) mRNA levels were significantly increased, while NF-κB p65 (pro-inflammatory) protein levels were decreased, compared to controls (Figures 3c to 3f). NF-κ p65 is a subunit of NF-κB, plays a critical role in the pathogenesis of multiple chronic inflammatory diseases by upregulating transactivation of inflammatory cytokines target genes ^20^; while IL-10 is an anti-inflammatory cytokine ^21^. Thus, valeric acid inhibited inflammatory responses of osteoclast-like cells and osteoblasts, which may explain its positive association with BMD, since inflammation may lead to bone loss ^19^, and the NF-κB can inhibit osteoblast functions ^22^.

## Discussion

We found that: **1)** GM biodiversity was negatively associated with forearm BMD; **2)** several individual bacterial species were significantly associated with BMD; **3)** *B.vulgatus* was negatively associated with BMD in ethnically distinct populations; **4)** several bacterial metabolic pathways were negatively associated with BMD; **5)** serum valeric acid levels were positively associated with L1-L4 BMD; **6)** *B.vulgatus* was negatively associated with serum valeric acid levels; and **7)** *B.vulgatus* may causally suppress valeric acid levels, presumably by inhibiting growth of valeric acid producing bacteria within the gut.

Subsequent *in vivo* and *in vitro* studies found that **1)** bone formation was decreased, and bone resorption increased, in OVX female mice fed *B.vulgatus*; **2)** valeric acid suppressed maturation of osteoclast-like cells, and promoted maturation of osteoblasts and extracellular matrix mineralization by osteoblasts *in vitro*; **3)** valeric acid treatment of osteoclast-like cells and osteoblasts *in vitro* suppressed NF-κB p65 protein production (pro-inflammatory), and enhanced IL-10 mRNA expression (anti-inflammatory). Thus, *B.vulgatus* appears to decrease BMD by reducing valeric acid production within the gut, resulting in enhanced inflammation and osteoclast activity, and reduced osteoblast activity.

The negative association between GM biodiversity and BMD in our discovery sample is consistent with previous reports. Wang *et al*. found elevated diversity in OP and osteopenia groups compared with controls ^8^, and Das *et al*. found a trend of higher Shannon index (α-diversity) in OP and osteopenia groups compared with controls (Fig 3D in ^6^) ^6^. Germ-free mice (no GM biodiversity exists) had higher bone mass than mice with normal GM ^5^, and that antibiotic treatment of mice early in life reduced GM biodiversity and increased bone size ^23^. Although GM biodiversity is generally believed to be beneficial ^24^, the above results suggest that the effects of GM biodiversity on health vary with the trait studied, environmental factors (e.g. diet and exercise) and individual species composition within GM.

In our study, *B.vulgatus* was negatively associated with L1-L4 BMD in the Chinese cohort (β = -0.027, *p*-value = 0.032), which has a consistent association with HTOT BMD in the US cohort (β = -0.018, *p*-value = 0.029). In fact, *B.vulgatus* tended to be consistently negatively (though non-significantly in many cases) associated with BMDs at all the measured various other skeletal sites in both of the two human cohorts (ED Table 8). To evaluate the overall effects of *B.vulgatus* on BMD, we further performed one-sided sign test ^25^ based on the coefficients of the regression of individual skeletal sites with *B.vulgatus* (ED Table 8). The null hypothesis (H0) is that regression coefficients are equal to zero. The alternative hypothesis (H1) is that regression coefficients are less than zero. The results indicated clearly that overall, there were significant negative associations between BMDs and *B.vulgatus* in both of the Chinese and the US cohorts (*p*-value_Chinese = 4.8 × 10^−7^, *p*-value_US = 9.8 × 10^−4^). In genomics studies, it is not uncommon to observe that a particular gene was significantly associated with different skeletal sites of BMDs in different populations. An example is given by the most prominent *WNT16* gene, which was found to be associated with BMD at different skeletal sites in different cohorts ^26^. The association and partial validation between *B.vulgatus* and BMD in the human cohorts was only mainly for discovery relative to the following-up functional validation studies. Following discoveries in humans, we subsequently validated the results by various *in vivo* and *in vitro* experiments. All of the results together furnish strong evidence for the significance of *B.vulgatus* on BMD and bone metabolism.

*B.vulgatus* is one of the most abundant bacteria in human gut ^27^, and can trigger proinflammatory NF-κB signaling pathways ^27^ associated with bone remodeling ^28^. To determine how GM influences BMD, we examined relationships between GM functional capacity and BMD variation. Most GM functional modules negatively associated with BMD (Table 2) involved GM metabolism. M00002, M00003 and M00855 involved glycolytic pathways or gluconeogenesis, both of which affect glucose metabolism, essential for cellular energy ^29^. Modules M00048 and M00050 both involve nucleic acid synthesis required for bacterial growth ^30, 31^. These collective findings demonstrate that human bone health is affected by GM, and that specific bacterial biosynthetic/metabolic pathways, contribute to these effects.

Since several GM functional modules were associated with BMD, and microbial metabolites contribute to host-microbiome interactions ^32^, we explored relationships between BMD and serum SCFAs. SCFAs are produced exclusively by GM ^15^, absorbed in the colon ^15^, and function as critical signalling molecules between the host and GM ^15^. Regarding the potential impact of SCFAs on bone health, butyrate has been reported to inhibit osteoclast differentiation and bone resorption in mice ^14, 33^. In the current study, we found that valeric acid (an SCFA) was positively associated with BMD, inhibited differentiation of osteoclast-like cells, and promoted differentiation and mineralization of osteoblasts. Valeric acid also suppressed NF-κB p65 protein production (pro-inflammatory), and enhanced expression of IL-10 mRNA (anti-inflammatory) by osteoclast-like cells and osteoblasts. This is consistent with previous reports that SCFAs reduced macrophage induced inflammation by suppressing NF-κB, and increasing IL-10, expression ^34^. IL-10 inhibits NF-κB production ^35^, and NF-κB promotes inflammation, which contributes to bone loss ^19^, and inhibits osteoblasts, which reduces new bone formation ^22^. These collective results are consistent with the conclusion that the protective effect of valeric acid on BMD is partially attributable to decreased osteoclast activity and increased osteoblast activity through increased IL-10 expression, and direct (via valeric acid) or indirect (via IL-10) inhibition of NF-κB expression ^28^. Valeric acid inhibits histone deacetylase (HDAC), an enzyme that is important for epigenomic regulation of gene expression ^36^. HDAC is implicated in the pathogenesis of a number of diseases (e.g., cancer, colitis, cardiovascular disease and neurodegeneration), and HDAC inhibitors are considered as potential therapeutic agents ^36^. In irradiated mice, valeric acid can protect hematogenic organs, improve gastrointestinal tract function and enhance intestinal epithelial integrity to elevate survival rate ^37^. Our study here for the first time suggest that valeric acid could be used to treat/prevent OP.

Since both *B.vulgatus* and valeric acid were associated with BMD, we sought to determine whether *B.vulgatus* regulated bone metabolism by decreasing valeric acid levels. Regression and MR analyses demonstrated that *B.vulgatus* was negatively associated with valeric acid, and may causally down-regulate valeric acid levels. We also observed negative correlations between *B.vulgatus* and common gut probiotic bacteria ^16, 17^. Bacterial fermentation product levels (e.g., valeric acid) depend on the relative abundances of different GM populations, and on competitive and cooperative interactions between different GM species ^38^. GM in humans is dominated by negative interactions, and the *Bacteroidales*, which includes *B.vulgatus*, commonly express molecular mechanisms for delivering toxic payloads to other bacteria ^39^. Consequently, we speculate that *B.vulgatus* competitively inhibits growth of valeric acid producing probiotic bacteria, thereby reducing valeric acid levels.

In this study, we pioneered an innovative and comprehensive multi-omics approach for discovery in a Chinese cohort, followed by *in vitro* and *in vivo* functional validation experiments plus replication in an independent cohort of US Caucasians. We, for the first time, successfully identified and validated individual gut bacterial species, and their derived metabolites, that were significantly associated with human BMD. These findings provide a potential novel target and pathway for OP prevention and treatment during the critical menopausal period in aging women.

There are several strengths of our study. First, to our knowledge, this is currently the largest metagenomics study directly testing associations between GM and human BMD. Second, we used stringent inclusion and exclusion criteria to ensure that subjects were relatively homogeneous for age, ovarian function and living environment, thereby minimizing potential confounding factors. Third, functional mechanisms contributing to associations between GM and BMD were identified by using an innovative multi-omics approach to generate a comprehensive understanding of crosstalk, interactions, and causal inference of the interactions, between GM and human BMD. Finally, we used both statistical and experimental evidences to demonstrate the effects of specific bacterial species and GM-derived metabolites on BMD variation and regulation, and the causal effects of specific gut bacterial species on GM-derived metabolites in human serum.

In conclusion, our study provides compelling evidence for associations between GM, SCFAs and human BMD. Importantly, we demonstrated that *B.vulgatus* appears to play a critical role in regulating bone metabolism, through its effects on valeric acid production. Valeric acid, in turn, promotes differentiation and mineralization of osteoblasts, and suppresses osteoclast differentiation and inflammatory responses by osteoblasts and osteoclast-like cells. Our findings provide novel insights into the pathophysiological mechanisms of OP from a novel perspective of human microbiota and their functional products, SCFAs, in human serum and suggested potential new biomarkers (e.g., valeric acid) and treatment targets (e.g., elimination of *B.vulgatus* from the gut; dietary supplementation with valeric acid) for OP prevention/intervention/treatment.

## Online Methods

### Subjects recruitment and sample preparation

This study was approved by the Third Affiliated Hospital of Southern Medical University (Guangzhou City, China). It was performed under the principle of the Helsinki Declaration II. 517 independent unrelated peri-/post-menopausal Chinese women were recruited. The inclusion criteria included **1)** aged 40 years or older, **2)** being in menopause stage, and **3)** had lived in Guangzhou City for at least three months. Menopause is marked by the cessation of menstruation, where peri-menopause is a transition phase beginning at a woman’s last menstrual cycle and continuing through the following 12 months without a menstrual cycle; once there are no menstrual cycles for 1 year, we term it post-menopause ^10^. Briefly, exclusion criteria included the use of antibiotics, oestrogens, or anticonvulsant medications which may affect gut microbiota (GM) composition and/or bone metabolism in the past three months, as well as other diseases that could lead to secondary osteoporosis (OP). A detailed list of exclusion criteria is shown in Extended Data (ED) Table 9. We obtained signed informed consent from each subject before enrolling them into this study. Each subject filled out a questionnaire that collected information on age, medical history, family history, physical activity, alcohol consumption, diet habits, smoking history, nutrition supplements, etc.

We used a GE Lunar dual energy X-ray absorptiometry (DXA, GE Healthcare, Madison, WI, USA, version 13.31.016) with a standard scan model to measure the bone mineral density (BMD) of each subject at various skeletal sites, including the lumbar spine (L1-L4), left total hip (HTOT), femoral neck (FN), ultra-distal radius and ulna (UD-RU). We performed DXA scanner calibration with a daily special phantom scan for quality assurance. The accuracy of various skeletal sites BMD measurement was assessed by the coefficient of variation (CV%) of spine BMD, which was 0.89%.

Blood and stool samples were collected from each subject. Blood samples were collected after an overnight fast for > 8 hours, and used for serum analysis and DNA extraction with the SolPure DNA Kit (Magen, Guangzhou, China). Each faecal sample was frozen at -80 °C within 30 min of sample procurement, and used for GM DNA extraction with the E.Z.N.A.® Stool DNA Kit (Omega, Norcross, GA, USA). We stored the serum, blood and GM DNA samples at -80 °C until further analyses.

### Metagenomic shotgun sequencing and annotation

#### DNA library construction

Shotgun metagenomic sequencing was performed by LC-Bio Technologies (Hangzhou) CO., LTD. (Hangzhou, China, www.lc-bio.com; this company owns an LC Sciences R&D department in Houston, TX, USA, www.lcsciences.com). We constructed a faecal DNA library, and used Hiseq 4000 (Illumina, San Diego, CA, USA) and PE150 strategy to conduct metagenomic shotgun sequencing. The relative abundance of unigenes for a sample was estimated by transcripts per kilobase million (TPM, Formula 1, where k was the k^th^ unigene, r was number of unigene reads, and L was unigene length) based on the number of aligned reads and the unigene length by Bowtie2 v2.2.0.

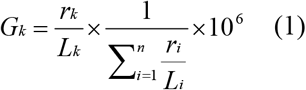

#### Taxonomic and functional annotation of unigenes

The unigenes were aligned against the NCBI NR database (ftp://ftp.ncbi.nlm.nih.gov/blast/db/FASTA/nr.gz) by DIAMOND software ^40^ with lowest common ancestor algorithm ^41^ for taxonomic annotation, and against protein reference based on the KEGG pathway dataset (https://www.genome.jp/kegg/pathway.html) for functional annotation. The relative abundance of each annotated functional module was the sum of relative abundance of the unigenes within this module. We chose the modules whose relative abundance > 0.10% to estimate the association between the KEGG modules and BMD variation by performing partial Spearman correlation analysis, and adjusted for a number of covariates, including age, body mass index (BMI), years since menopause (YSM), follicle stimulating hormone (FSH), exercise, and family annual income. Results with *p*-values < 0.01 were considered statistically significant.

Detailed information about faecal DNA library construction, raw data preprocessing and cleaning, taxonomic and functional annotation of unigenes, was shown in supplementary information.

### Unigene accumulation curve construction and species biodiversity calculation

We first counted the number of unigenes at different sample sizes. The R package “ggplot2” was used to generate a unigene accumulation curve. We plotted the number of unigenes (Y-axis) against the number of samples (X-axis).

The α-diversity of GM was estimated based on the species profile of each sample according to the Shannon index by using R package “vegan”. Then, partial Spearman correlation analysis was performed to estimate the association between the Shannon index and BMD variation. The association between bacterial species profiles and BMD variation was calculated by the program Microbiome Regression-based Kernel Association Test (MiRKAT) ^12^, by which we first calculated different kinds of kernels (including weighted and unweighted UniFrac distance matrices and Bray-Curtis distance), and then obtained the optimal kernel to estimate the association between GM species profiles and BMD variations. In these analyses, we adjusted the same set of covariates as the KEGG modules analysis mentioned above. Results with *p*-values less than 0.05 were considered statistically significant.

### Measurement of short chain fatty acids (SCFAs)

2 μl of the supernatants extracted from serum samples were analyzed with gas chromatography-tandem mass spectrometry (GC-MS/MS, 7890B-7000D, Agilent Technologies Inc, Santa Clara, CA, USA) by Wuhan Metware Biotechnology Co., Ltd (Wuhan City, China, www.metware.cn) based on a fused silica capillary column (DB-FFAP, 30 m × 0.25 mm × 0.25 µm, Agilent Technologies Inc, Santa Clara, CA, USA). Then we constructed standard curves to calculate concentration of SCFAs. Detailed conditions of GC-MS/MS, quality control (QC), preparation of SCFAs calibration standards, and SCFAs concentration calculation by standard curves, were shown in supplementary information.

### Association analysis among GM, SCFA, and BMD

We treated BMD as a continuous variable and then explored the association among GM, SCFA, and BMD by using different association analysis methods. In this study, we focused on non-rare species (relative abundance > 0.10%) because rare species typically contribute significantly less to functional diversity than non-rare species due to their low abundances ^42^; meanwhile, the rare species are generally difficult to cultivate ^42^, which might make our subsequent experimental validation *in vivo* difficult.

For the association analysis, first, we used the “igraph” package and the “psych” package of R software to identify correlations between bacterial species. Spearman correlation analysis was performed on the relative abundance of each bacterial species. Meanwhile, we used the same method to identify correlations between specific bacterial species and probiotic bacteria reported in two previous reviews which summarized the most notable probiotics ^16, 17^. The reported probiotic bacteria were pooled for such analyses due to the small relative abundances of each of the individual species (less than 0.10%). Furthermore, the R package “pheatmap” package was used to create a heatmap for visualization of the bacterial species correlation.

Second, for those highly correlated bacterial species (with correlation coefficients [γ’s] ≥ 0.80) that might cause a multiple co-linearity problem, we retained the bacterial species with a higher relative abundance and removed others as bacteria with relatively higher relative abundance may render higher power in association testing. Constrained linear regression analysis ^13^ was then performed by using Stata 14 software with the “cnsreg” function, in which BMD variation was considered as the dependent variable, and the relative abundance of bacterial species was set as the independent variable. A number of covariates were adjusted in the regression analysis, including age, BMI, YSM, FSH, exercise, and family annual income. The threshold of exercise was 2.5 hours per week based on a published guideline ^43^. The family annual income was separated according to the local economic report. As microbiome data are often obtained at different taxonomic ranks (including species, genus, family, order, class and phylum), we performed subcomposition analysis using previously published protocols ^13^, and converted the bacterial species relative abundance data into subcompositions within each phylum by centered log ratio transformation, to find out which bacterial species within a given phylum is associated with BMD variation. The formula of constrained linear regression analysis is shown below (Formula 2), where p was the phylum level, n was the number of phyla, s was species level, and m was the number of bacterial species. Bacterial species with a *p*-value of the regression coefficient (β) less than 0.05 were defined as BMD-associated bacterial species.

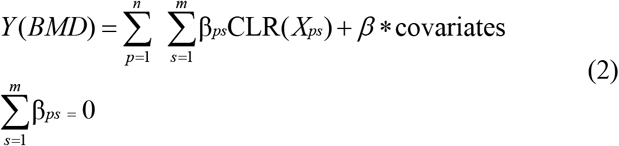

Third, we used multiple linear regression analysis to identify BMD-associated SCFAs (Formula 3), where we performed log transformation for the SCFA concentration because of their non-normalization distribution and adjusted the same set of covariates mentioned above.

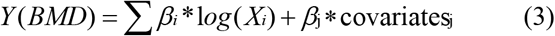

Finally, we performed constrained linear regression analysis (Formula 4) to identify SCFA-associated bacterial species, while adjusting for age, BMI, YSM, and FSH as covariates.

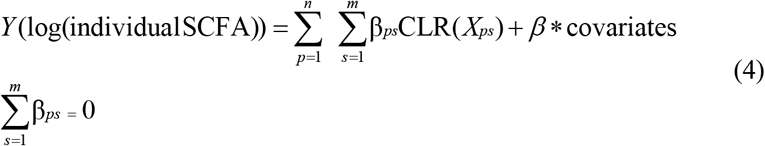

#### Replication in the Caucasian cohort

A total of 59 Caucasian females, aged 60 years or older, were recruited from the ongoing Louisiana Osteoporosis Study (from 2011 to current), which aims to build a large sample pool in greater New Orleans and surrounding areas in Southern Louisiana, USA for investigating genetic and environmental factors for musculoskeletal disorders ^44^. This study was approved by the institutional review board of Tulane University (New Orleans, LA, USA), and a written consent form was signed by each participant before data and bio-sample collection. Individuals who had pathological conditions that may influence BMD (e.g., a bilateral oophorectomy, chronic renal failure, liver failure, lung disease, gastrointestinal diseases, and inherited bone diseases), or may influence GM (e.g., taking antibiotics, having gastroenteritis, major surgery involving hospitalization, and inter-continental travel in the past three months) were excluded. For each participant, BMD was measured by using a Hologic Discovery-A DXA machine (Hologic Inc., Bedford, MA, USA) and information on age, medical history, physical activity, alcohol consumption, diet habits, smoking history, and nutrition supplements was assessed by a questionnaire. The accuracy of BMD measurement as assessed by the CV% of L1-L4 BMD was 0.54%. The shotgun metagenomics sequencing on the collected faecal samples were performed by Alkek Center for Metagenomics and Microbiome Research (CMMR), Baylor College of Medicine. We conducted partial Spearman correlation analysis for the identification of correlation between GM biodiversity and BMD variation, and we also used constrained linear regression analysis (Formula 2), as described above, to identify BMD-associated individual bacterial species. The covariates included age, BMI, exercise, and bone fracture.

### *In vivo* mouse experiment with *B.vulgatus*

#### Ovariectomy (OVX) and B.vulgatus gavage

All the procedures involving mice were approved by the Southern Medical University Animal Management Committee (Guangzhou City, China). Seven-week-old female C57BL/6 mice (specific-pathogen free grade) were purchased from the Laboratory Animal Center of the Southern Medical University (Guangzhou, China). The mice were fed freely with food and water for one week to acclimate to the new environment before experiments. Then bilateral OVX surgery was performed with general anaesthesia by the dorsal approach because eight-week-old C57BL/6J mice are suitable animal model for postmenopausal osteoporosis ^45^. After one week for recovery, the mice were separated randomly into two groups (9 samples/group): Group I mice were gavaged with 100 μl of *B.vulgatus* (ATCC8482, 5 × 10^8^ colony-forming units/mouse, ATCC, Manassas, VA, USA; lot number: 70009621), while Group II mice were gavaged with 100 μl of normal saline (NS). *B.vulgatus* was cultivated according to the product description and harvested in log phase for gavage. All mice were gavaged every other day and continued to feed freely with common food and water under the same environmental conditions. After gavaging for eight weeks, the mice were sacrificed and used for blood and bone measurements.

#### BMD measurement and bone morphology evaluation by micro-computed tomography (μCT)

We used LaTheta LCT 200 μCT (Aloka, Japan) to measure the BMD of the 5^th^ lumbar vertebral body. The instrument accuracy was 1 ± 0.005 mg/cm^3^. We performed phantom scan at first for QC. Then we scanned the lumbar spine to obtain lumbar vertebral body BMD. Meanwhile, for visualization and comparison of bone microstructure changes, we randomly selected a bone sample (the 5^th^ lumbar vertebrae) from each of the two mice groups (Group I with *B.vulgatus* gavage and Group II with NS gavage), and used μCT100 (Scanco Medical AG, Bassersdorf, Switzerland) with a resolution of 7.4 μm for three-dimensional reconstruction.

#### Analysis of serum biomarkers

We used ELISA kits to measure osteocalcin (Cloud-Clone Corp, Katy, TX, USA) and C-telopeptide of type I collagen (Cusabio, Houston, TX, USA), which are markers of bone formation and bone resorption, respectively.

#### Histomorphometry

We used hematoxylin-eosin (ZSGB-BIO, Beijing, China) staining to assess histomorphology of trabecular bone in the 5^th^ lumbar vertebrae.

#### Statistical analysis

We used two-sample t-test to identify whether BMD and serum biomarkers were significantly different between the two groups.

Details about scanning parameters of μCT and histomorphometry process were shown in supplementary information.

### Mendelian randomization (MR) analysis for BMD-associated GM and SCFA

#### Whole genome sequencing (WGS)

WGS was performed by BGI Genomics Co. Ltd (Shenzhen, China; https://www.genomics.cn/) with BGISEQ-500 platform.

Details including quality control were shown in supplementary information.

#### Genome-wide association study (GWAS)

We used PLINK 1.9 software with the default criteria as follows to identify *B.vulgatus* and valeric acid-associated SNPs: the SNPs with missing call rate < 0.1, minor allele frequencies > 0.01, and Hardy-Weinberg equilibrium *p*-value > 1.0 × 10^−5^ were included for subsequent analyses.

#### MR analysis

Based on *B.vulgatus* and valeric acid GWAS results, we investigated the causality of *B.vulgatus* exposure on valeric acid outcome with one-sample MR analysis, which is a commonly used method. One-sample MR analysis uses genetic variants, exposure, and outcome from the same individua, which provides greater control of the analysis (such as determining exposure category - a continuum or in categories, examining associations between genetic instruments and confounders) than two-sample MR analysis ^46^. For the implementation of MR, we first selected independent genetic variants (r^2^ ≤ 0.001) associated with the *B.vulgatus* (*p-*values < 1 × 10^−5^) as the instrumental variables (IVs) as done earlier ^47^. We then obtained the corresponding effect estimates of these SNPs from the valeric acid GWAS analysis. When the target SNPs were not available in the valeric acid GWAS result, we used proxy SNPs that were in high linkage disequilibrium (LD, r^2^ > 0.80) with the SNPs of interest. The proxy SNPs were identified based on 1000 genomes project data ^48^. To ensure the SNPs used as IVs for *B.vulgatus* are not in LD with one aother, a vital assumption of MR, we calculated pairwise-LD between all of our selected SNPs in the 1000 Genomes European reference sample using PLINK 1.9. For all the pairs of SNPs determined to violate the independence assumption with r^2^ > 0.01, we retained only the SNPs with the smaller *B.vulgatus* association *p*-values. Overall, we obtained 15 LD-independent SNPs that achieved genome-wide significance for *B.vulgatus* and valeric acid. Therefore, these 15 SNPs remained for performing the MR analysis (Extended Data Table 10). Weighted median method, maximum likelihood estimation (MaxLik) and inverse-variance weighted (IVW) ^49^ were performed to identify causal relationships between *B.vulgatus* and valeric acid by weighting the effect estimate of each SNP on valeric acid with its effect on *B.vulgatus*. Because SCFA was only produced from GM ^14^, we only chose single direction MR (*B.vulgatus* as exposure and valeric acid as outcome) rather than bidirectional MR analysis. Because the presence of horizontal pleiotropy could bias the MR estimates, we used the intercept that deviates from the MR-Egger method to examine the presence of horizontal pleiotropy as described in the previous study ^47^.

### Effects of valeric acid on osteoclastogenesis and osteoblastogenesis

#### In vitro differentiation of osteoclast-like cells

We used the murine monocyte/macrophage cell line RAW264.7 (Chinese Academy of Sciences Cell Bank, Shanghai, China) as osteoclast precursors, which can differentiate into osteoclast-like cells in the presence of receptor activator of nuclear factor-κB ligand (RANKL, 50 ng/mL, PEPROTECH, Rocky Hill, NJ, USA). The cells were grown in Dulbecco’s Modified Eagle Medium (DMEM, GIBCO/Invitrogen, Carlsbad, CA, USA), containing 10% fetal bovine serum (FBS, GIBCO/Invitrogen, Carlsbad, CA, USA), 100 U/ml penicillin and 100 mg/ml streptomycin sulfate (GIBCO/Invitrogen, Carlsbad, CA, USA), at 37°C in a humidified atmosphere of 95% air and 5% CO_2_, and treated with various concentrations (0, 0.001, 0.01, 0.1, 1 mM) of valeric acid (TCI, Tokyo, Japan). We changed the medium every other day, and fixed the cells at the end of the 5^th^ day. Then we performed tartrate-resistant acid phosphatase (TRAP) staining (Solarbio, Beijing, China) to observe TRAP-positive multinucleated (≥ 3 nuclei) cells.

#### In vitro differentiation of osteoblasts

MC3T3-E1 cells (Chinese Academy of Sciences Cell Bank, Shanghai, China) were maintained in osteoblastogenic medium (AAPR99-C500, Pythonbio, Guangzhou, China) with 10% FBS, 100 U/ml penicillin, and 100 mg/ml streptomycin sulfate at 37°C with 5% CO2, and treated with various concentrations of valeric acid (0, 0.001, 0.01, 0.1, 1 mM) for 14 days. Then alkaline phosphatase (Beyotime, Shanghai, China) staining and alizarin red S (Solarbio, Beijing, China) staining were performed according to standard techniques to evaluate osteoblast differentiation and extracellular matrix mineralization, respectively.

### Gene expression in osteoclast-like cells and osteoblasts

Total protein and RNA were extracted from osteoclast-like cells and osteoblasts to examine expression of the selected genes. Protein concentration was determined by the BCA protein assay kit (Thermo scientific, Rockford, IL, USA). Since NF-κB signaling pathway was reported to activate osteoclastogenesis ^28^ and inhibit osteoblastogenesis ^22^, and SCFAs can suppress this pathway ^34^, we explored whether the suppressive effect and stimulating effect of valeric acid on osteoclast-like cells and osteoblasts, respectively, were at least partially through inhibition of the NF-κB signaling pathway. Western blot (WB) analysis was performed with NF-κB signaling pathway-associated primary antibody targeting NF-κB p65 (Bclonal, Beijing, China) in osteoclast-like cells and osteoblasts, and GAPDH (Proteintech, Chicago, IL, USA) was used as a reference protein. After validating the association between valeric acid and the NF-κB signaling pathway, we further explored whether IL-10 in osteoclast-like cells and osteoblasts contributed to this association. We examined IL-10 because it can be increased by SCFAs ^34^ and inhibit activation of NF-κB ^35^. Real-time PCR analysis (SYBR green real time PCR master mix, TOYOBO, Osaka, Japan) was performed for the transcriptional levels of IL-10. Relative expression of IL-10 was normalized to mouse-18S expression, and calculated using the comparative threshold cycle (Ct) method ^50^. The primers of IL-10 were GGAGCTGAGGGTGAAGTTTGA (sense) and GACACAGACTGGCAGCCAAA (antisense). The primers of mouse-18S were CGATCCGAGGGCCTCACTA (sense) and AGTCCCTGCCCTTTGTACACA (antisense).

### Online resources and abbreviations

Online resources and abbreviations are provided in supplementary information.

## Data Availability

The data that support the findings of this study are available from the corresponding author upon request and approval of the team and respective institutions.

## Acknowledgement

HW Deng and H Shen were partially supported by grants from the National Institutes of Health [U19AG05537301, R01AR069055, P20GM109036, R01MH104680, R01AG061917], and the Edward G. Schlieder Endowment and the Drs. W. C. Tsai and P. T. Kung Professorship from Tulane University. J Shen was partially supported by grants from the Science and Technology Program of Guangzhou, China [201604020007], and the National Natural Science Foundation of China [81770878]. HM Xiao was partially supported by the National Key R&D Program of China (2016YFC1201805 and 2017YFC1001100).

We thank Feng-Zhu Sun, Guo-Zhen Wang, Jun Wang, Wei-Chang Huang, Zun Wang, Yong Liu, Lin-Dong Jiang, Rui-Ke Liu, Zhi-Mei Feng, Yuan-Yuan Hu, Lin-Ping Peng and Chun-Ping Zeng for their suggestions and supports for this study.

## Author contributions

HWD conceived, designed, initiated and directed the whole project. JG, QZ, KJS, XHM, FYL, and CLG contributed to the data analysis. JS and HMX managed the study done in their institutions. DYP, ZC, and ZFL performed clinical diagnosis and recruited subjects. CP, XJX, YCC, RZ, XFW, ZXA, JML, YQS, and YHZ collected samples and clinical phenotypes. SJY conducted animal experiments. XL drafted the manuscript. HWD revised, rewrote/re-structured some sections and finalized the manuscript. NJY, QZ, BYG, HML, WQL, CJP, XMS and HS contributed to text revision and/or discussion.

## Author information

The authors declare no competing financial interests.

Correspondence and requests for materials should be addressed to Hong-Wen Deng, Jie Shen and Hong-Mei Xiao.

